# The impact of BNT162b2 mRNA vaccine on adaptive and innate immune responses

**DOI:** 10.1101/2021.05.03.21256520

**Authors:** Konstantin Föhse, Büsra Geckin, Martijn Zoodsma, Gizem Kilic, Zhaoli Liu, Rutger J. Röring, Gijs J. Overheul, Josephine S. van de Maat, Ozlem Bulut, Jacobien J. Hoogerwerf, Jaap ten Oever, Elles Simonetti, Heiner Schaal, Ortwin Adams, Lisa Müller, Philipp Niklas Ostermann, Frank L. van de Veerdonk, Leo A.B. Joosten, Bart L. Haagmans, Reinout van Crevel, Ronald P. van Rij, Corine GeurtsvanKessel, Marien I. de Jonge, Yang Li, Jorge Domínguez-Andrés, Mihai G. Netea

**Affiliations:** Department of Internal Medicine and Radboud Center for Infectious Diseases, Radboud University Medical Center, Nijmegen, The Netherlands; Radboud Institute for Molecular Life Sciences, Radboud University Medical Center, Nijmegen, The Netherlands; Department of Computational Biology for Individualised Infection Medicine, Centre for Individualised Infection Medicine (CiiM), a joint venture between the Helmholtz-Centre for Infection Research (HZI) and the Hannover Medical School (MHH), Hannover, Germany; TWINCORE, a joint venture between the Helmholtz-Centre for Infection Research (HZI) and the Hannover Medical School (MHH), Hannover, Germany; Department of Medical Microbiology, Radboud University Medical Center, Nijmegen, The Netherlands; Department of Laboratory Medicine, Laboratory of Medical Immunology, Radboud Center for Infectious Diseases, Radboudumc, Nijmegen, The Netherlands; Institute of Virology, Medical Faculty, University Hospital Düsseldorf, Heinrich-Heine-Universität, Düsseldorf, Germany; Department of Medical Genetics, Iuliu Hatieganu University of Medicine and Pharmacy, Cluj-Napoca, Romania; Department of Viroscience, Erasmus MC, Rotterdam, The Netherlands

## Abstract

The mRNA-based BNT162b2 protects against severe disease and mortality caused by SARS-CoV-2 through induction of specific antibody and T-cell responses. Much less is known about its broad effects on immune responses against other pathogens. In the present study, we investigated the specific adaptive immune responses induced by BNT162b2 vaccination against various SARS-CoV-2 variants, as well as its effects on the responsiveness of human immune cells upon stimulation with heterologous viral, bacterial, and fungal pathogens. BNT162b2 vaccination induced effective humoral and cellular immunity against SARS-CoV-2 that started to wane after six months. We also observed long-term transcriptional changes in immune cells after vaccination, as assessed by RNA sequencing. Additionally, vaccination with BNT162b2 modulated innate immune responses as measured by the production of inflammatory cytokines when stimulated with various microbial stimuli other than SARS-CoV-2, including higher IL-1/IL-6 release and decreased production of IFN-α. Altogether, these data expand our knowledge regarding the overall immunological effects of this new class of vaccines and underline the need of additional studies to elucidate their effects on both innate and adaptive immune responses.

## Introduction

The mRNA vaccine developed by BioNTech and Pfizer (BNT162b2) was approved for emergency use due to the protection induced against SARS-CoV-2 infection. It took less than eight months after trials started to achieve this landmark. This vaccine is based on a lipid nanoparticle–formulated, nucleoside-modified mRNA that encodes a prefusion stabilized form of the spike (S)-protein derived from the SARS-CoV-2 strain isolated at the beginning of the outbreak in Wuhan, China [1]. Multiple phase-3 trials have demonstrated that BNT162b2 elicits broad humoral and cellular-specific responses, providing protection against COVID-19 [1–3].

While the induction of specific immunity against SARS-CoV-2 has been intensively studied, much less is known about the effects of this new class of mRNA vaccines against heterologous pathogens. The lipid nanoparticle (LNP) component of those vaccines was reported to induce strong pro-inflammatory responses [4], and recent studies have shown that BNT162b2 can also induce long-term transcriptional changes in myeloid cells [5,6]. This suggests that the response of immune cells against various microorganisms other than SARS-CoV-2 could also change after BNT162b2 vaccination. Other vaccines, such as Bacillus Calmette-Guérin (BCG) or influenza A virus vaccines, but also the novel adenoviral-based COVID-19 vaccines, have been shown to induce long-term functional changes in innate immune cells, also called *trained immunity*, that subsequently results in heterologous protective effects [7–9]. There is much less data available on the functional effects of BNT162b2 vaccination on immune responses towards other pathogens than SARS-CoV-2.

With this in mind, we investigated the effects of BNT162b2 vaccination on both the specific adaptive immune responses and the responsiveness of human immune cells upon stimulation with heterologous pathogens. These experiments confirmed that BNT162b2 vaccination of healthy individuals induced effective humoral and cellular immunity against SARS-CoV-2, which started to wane after six months, especially against new variants. Interestingly, RNA sequencing revealed long-term changes in the transcriptional programs of immune cells after administration of the BNT162b2 vaccine, and vaccination also modulated the production of inflammatory cytokines upon stimulation with viral, bacterial, and fungal stimuli. The synthesis and release of myeloid-derived cytokines from the IL-1/IL-6 pathway tended to be higher six months after the first dose of BNT162b2. In contrast, the production of IFN-α after stimulation with SARS-CoV-2, TLR3 ligand poly I:C, and TLR7/8 ligand R848 decreased after vaccination. Altogether, we observed that administration of the BNT162b2 vaccine modulated innate immune responses up to one year after the initial vaccination. These data contribute to our understanding of the broad immunological effects of mRNA vaccines and underline the importance of performing additional studies to elucidate their full potential effects on innate and adaptive immune responses.

## Results

### Participants and study design

Sixteen healthcare volunteers who received the BNT162b2 mRNA COVID-19 vaccine as per national vaccination campaign were initially recruited in the study. Participants were 26-59 years of age (mean age 39.31±11.3 years), 7 men and 9 women, and without known acute or chronic diseases.

Samples were collected at five time points in accordance with the phase 1 trial performed by BioNTech and Pfizer [1]: before vaccination (t0), three weeks after the first dose of 30 μg of BNT162b2 (t1), two weeks after the second dose (t2) – i.e. 5 weeks after the first dose, six months after the first dose (t3), and four weeks after the booster vaccination, which was approximately one year after the first dose (t4) to obtain a broader view on potential long-term effects of the vaccination. The study design is shown in Fig 1.

**Fig 1.**
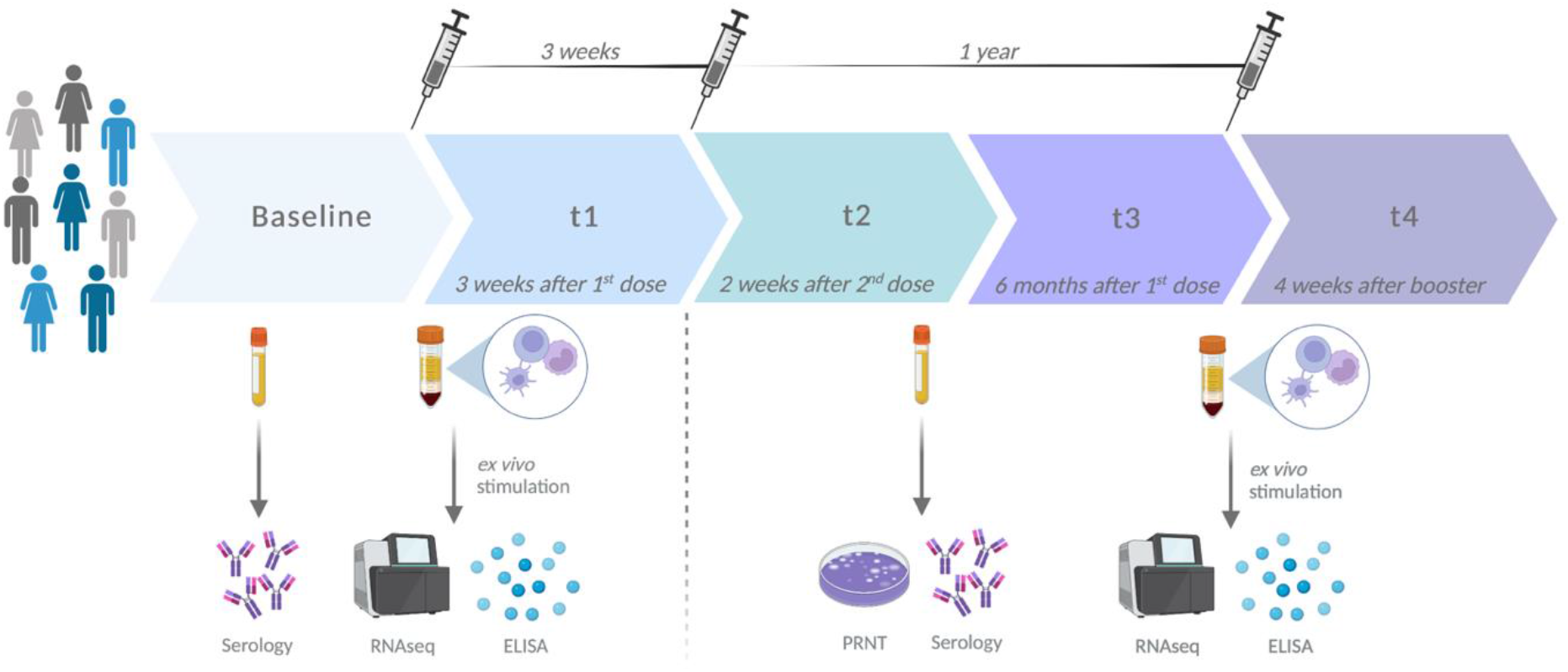
Study design. Created with BioRender.com.

### Long-term antibody concentrations and neutralization capacity against SARS-CoV-2 variants after vaccination with BNT162b2

First, we examined the concentration of receptor-binding domain (RBD)- and spike protein (S)-binding antibody isotype concentrations at given time points (Fig 2a). BNT162b2 vaccination elicited high IgG anti-S and anti-RBD concentrations already after the first vaccination and even stronger responses after the second dose of the vaccine. The antibody concentrations significantly decreased six months after vaccination and rose back to the levels of t2 after the booster vaccination.

**Fig 2.**
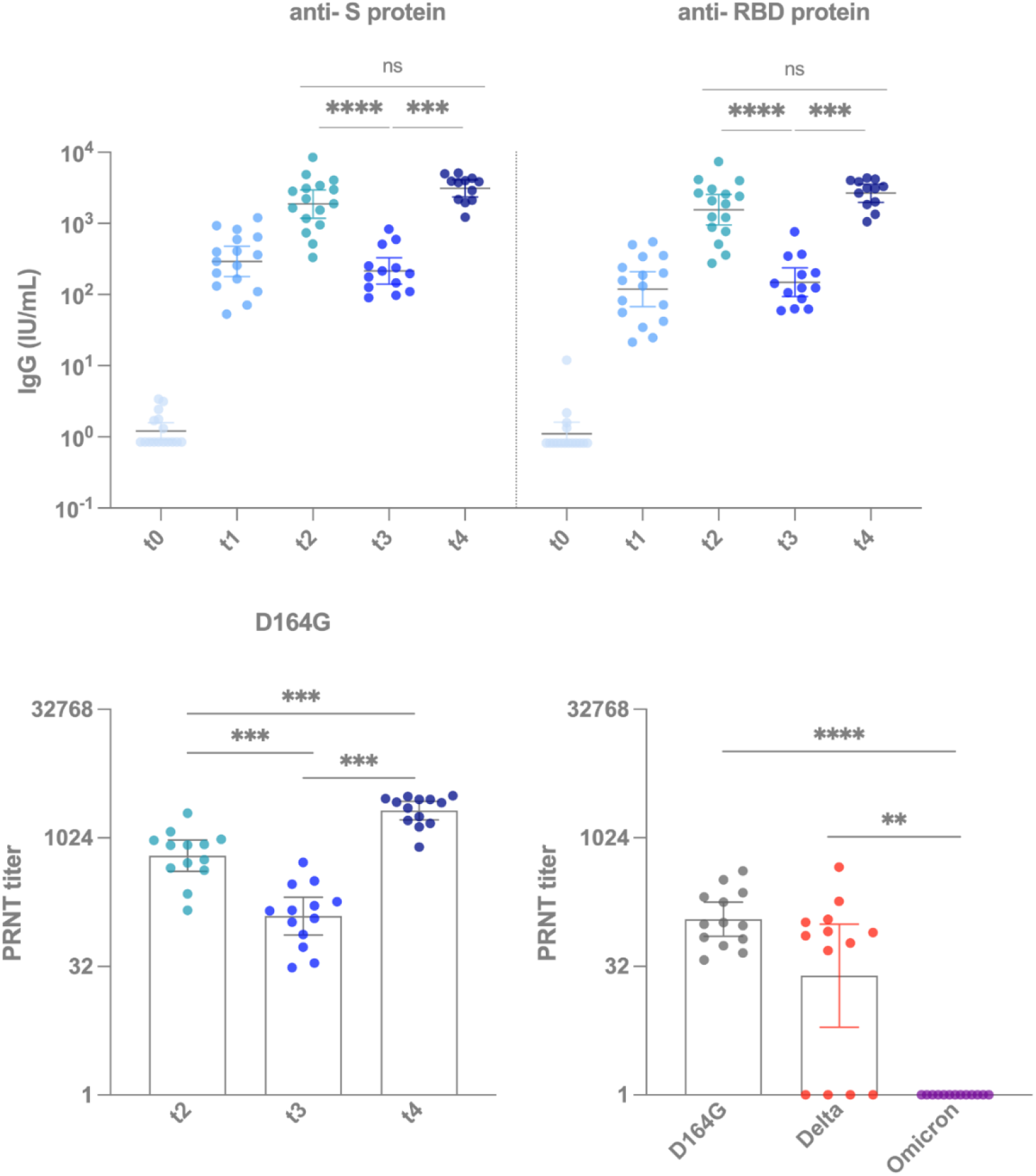
BNT162b2 vaccination induced effective antibody responses, but neutralizing capacity started to wane after six months. **(a)** S- and RBD-protein specific IgG measured from participants’ plasma collected at t0 (before vaccination), t1 (three weeks after the first dose), t2 (two weeks after the second dose), t3 (six months after the first dose) and t4 (four weeks after the booster vaccination, which was approximately one year after the first dose). Wilcoxon test (not corrected for multiple comparisons) is used to compare different time points. **(b)** PRNT from sera of the participants on Wuhan Hu-1 D164G after vaccination at depicted time points. Wilcoxon paired test (not corrected for multiple comparisons) is used to compare two time points to each other. **(c)** PRNT from sera of the participants on different variants six months after the first dose. Kruskal-Wallis test followed by Dunn’s multiple comparison is used to compare different variants. * p<0.05, ** p<0.01, *** p<0.001, **** p<0.0001. S protein, spike protein; RBD protein, receptor binding domain protein; IgG, immunoglobuline G; PRNT, plaque reduction neutralisation test.

To investigate the neutralizing capacity of the serum against SARS-CoV-2 variants, we performed 50% plaque reduction neutralization testing (PRNT50) using sera collected at t2, t3, and t4 (Fig 2b). After the second dose of BNT162b2, all the serum samples neutralized the D614G strain with titers of at least 1:146, and the geometric mean neutralizing titer (GMT) was 454 IU/ml. Similar to the antibody concentrations, the neutralizing capacity dropped significantly after six months to a GMT of 89 IU/ml. Booster vaccination led to a significant increase in neutralizing titers at t4 (GMT 1533 IU/mL) compared to t2. Finally, we analyzed the neutralizing capacity against different variants at t3. We observed that four of the thirteen samples (31%) failed to neutralize the delta variant, and none were able to neutralize the omicron variant (Fig 2c).

### Vaccination with BNT162b2 induces long-term transcriptional changes in immune cells

We assessed the potential effects of BNT162b2 vaccination on the transcriptional activity of immune cells using bulk RNA sequencing after *ex vivo* stimulation of peripheral blood mononuclear cells (PBMCs) obtained from healthy volunteers before and after vaccination. PBMCs were stimulated with heat-inactivated SARS-CoV-2, as well as with the heterologous stimuli R848, heat-inactivated influenza H1N1 virus or culture medium (RPMI1640; used as an unstimulated control condition).

First, we analyzed the responsiveness of the cells by comparing the number of differentially expressed genes (DEGs) induced by specific and non-specific viral stimuli to RPMI treatment within each time point (Figs 3a and 3b). The number of DEGs in PBMCs in response to heat-inactivated influenza, SARS-CoV-2, and R848 stimulation decreased after vaccination compared to RPMI. Especially after the stimulation of PBMCs with SARS-CoV-2, the number of DEGs was substantially lower at the late time points after BNT162b2 immunization. These results show that BNT162b2 vaccination notably affects transcriptional responses to SARS-CoV-2 and heterologous stimuli in PBMCs: interestingly, cells seem to respond less strongly to various stimulations when isolated from volunteers after BNT162b2 vaccination.

**Fig 3.**
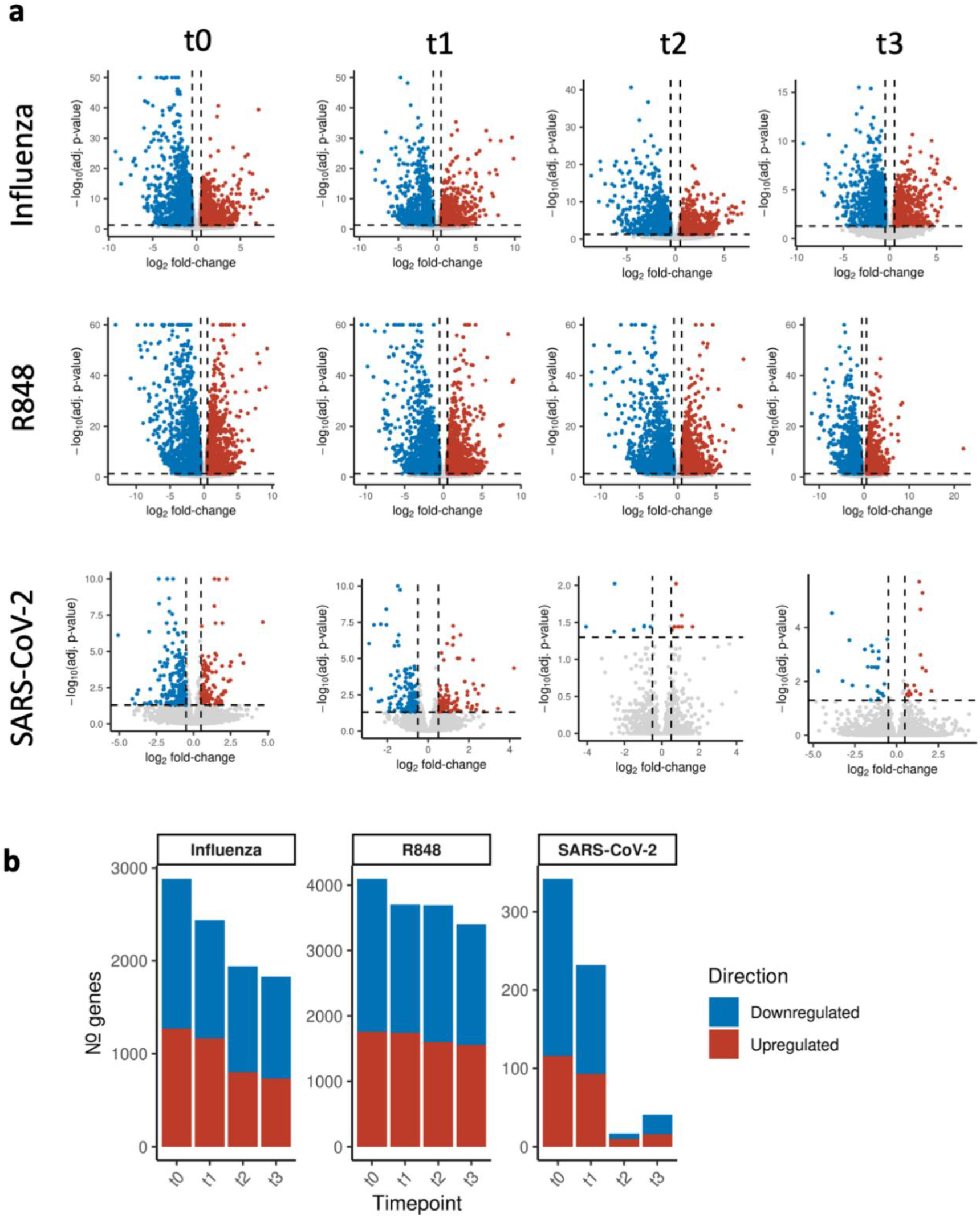
Responsiveness to viral stimuli compared to untreated RPMI cells decrease following BNT162b2 vaccination. **(a)** Volcano plots presenting the DEGs, showing the log2-fold-change (x-axis) vs. the negative log10 of the p-value (y-axis), following the stimulation of the cells (with influenza, R848 or SARS-CoV-2) and RPMI at t0 (before vaccination), t1 (three weeks after the first dose), t2 (two weeks after the second dose) and t3 (six months after the first dose). **(b)** Barplot depicting the total number of DEGs in response to stimulation with influenza, R848 or SARS-CoV-2 compared to RPMI at t0, t1, t2 and t3. DEGs, differentially expressed genes.

Gene set enrichment was performed to analyze the main cellular processes and pathways affected by the vaccination. Compared to RPMI, chemotaxis pathways were upregulated in all stimulated cells at each time point, whereas the upregulation of T-cell activation and signaling showed a varying pattern (S Fig 1). Interestingly, BNT162b2 vaccination first dampened T cell activation in SARS-CoV-2 stimulated cells, and after six months, the effect reversed. Stimulation with the TLR7/8 agonist R848 induced especially expression of genes related to cell cycle. Conversely, we observed a general downregulation of the gene expression of pathways associated with monocyte and dendritic cell activation. Genes related to (type I) IFN responses and innate antiviral responses were only downregulated following stimulation with influenza, and this was not affected by vaccination. On the contrary, BNT162b2 vaccination induced a persistent downregulation in pathways associated with inflammatory responses, while this effect occurred in the SARS-CoV-2-stimulated cells only after six months.

Next, we compared the transcriptional activity of the PBMCs at each time point to pre-vaccination levels within each stimulation to determine if the vaccine induces memory against a particular stimulus (Fig 4a). The number of DEGs in PBMCs incubated with influenza, R848 and in RPMI cells increased marginally with each time point, with R848 stimulation showing the most notable change in the numbers. Stimulation with SARS-CoV-2 showed a contrasting pattern with an increased number of DEGs after primary vaccination that returned to baseline levels at the following time points.

**Figure 4.**
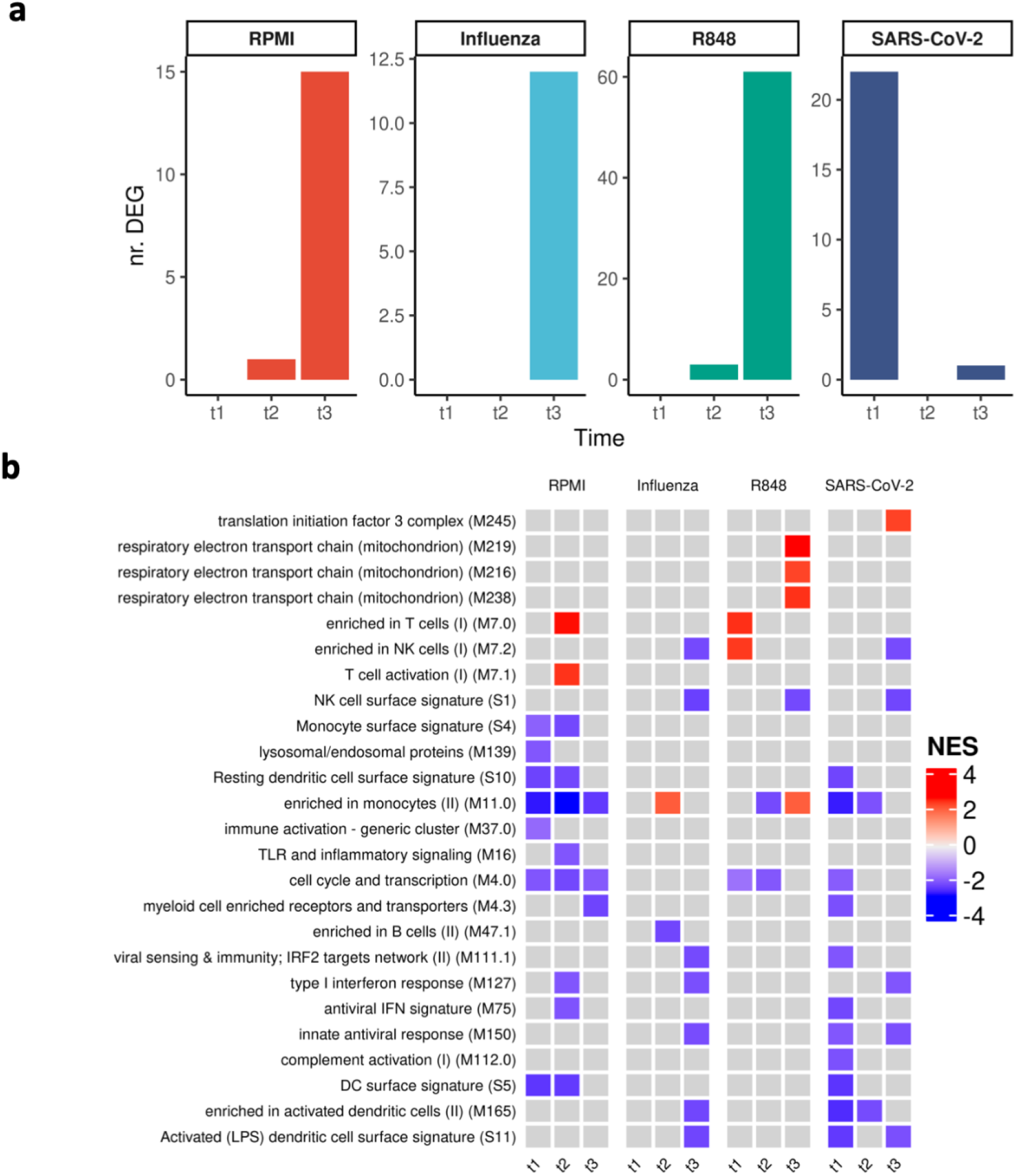
DEGs show variation in the course of vaccination and pathways are mostly downregulated compared to the situation before vaccination. **(a)** Barplot showing the number of DEGs of stimulated cells with RPMI, influenza, R848 and SARS-CoV-2 at t1 (three weeks after the first dose), t2 (two weeks after the second dose) and t3 (six months after the first dose) compared to t0 (before vaccination). **(b)** Heatmap depicting gene set enrichment of cells stimulated with RPMI, influenza, R848 and SARS-CoV-2 at t1, t2 and t3 to t0. Colors indicate correlation coefficients from negative (blue) to positive (red). Pathways with p adj < 0.0001 are shown. DEGs, differentially expressed genes.

Gene set enrichment analysis within each stimulus showed downregulation in the type I interferon pathways compared to baseline at t2 and t3 for all stimuli except R848 (Fig 4b). Generally, antiviral immune response-related pathways were downregulated at different points after vaccination. Moreover, genes related to the enrichment of T cells were upregulated in RPMI-treated and R848-stimulated cells after vaccination.

### Vaccination with BNT162b2 modifies cytokine production by PBMCs in response to different stimuli

In parallel to assessing the transcriptional responses, we measured cytokine secretion by enzyme-linked immunosorbent assay (ELISA) following an *ex vivo* challenge with heterologous stimuli to understand the functional effect of the vaccine on the inflammatory response of PBMCs. To examine the dynamics of cytokine production, we measured IL-6, IL-1β, TNF-α, and IL-1Ra in response to SARS-CoV-2, the bacterial pathogen *Staphylococcus aureus*, and the fungal pathogen *Candida albicans* (Fig 5). IL-6, IL-1β, and IL-1Ra production capacity varied between time points, but tended to increase six months after the first dose of BNT162b2 compared to baseline. In contrast, the production of TNF-α did not differ before and after the vaccination (S Fig 2).

**Figure 5.**
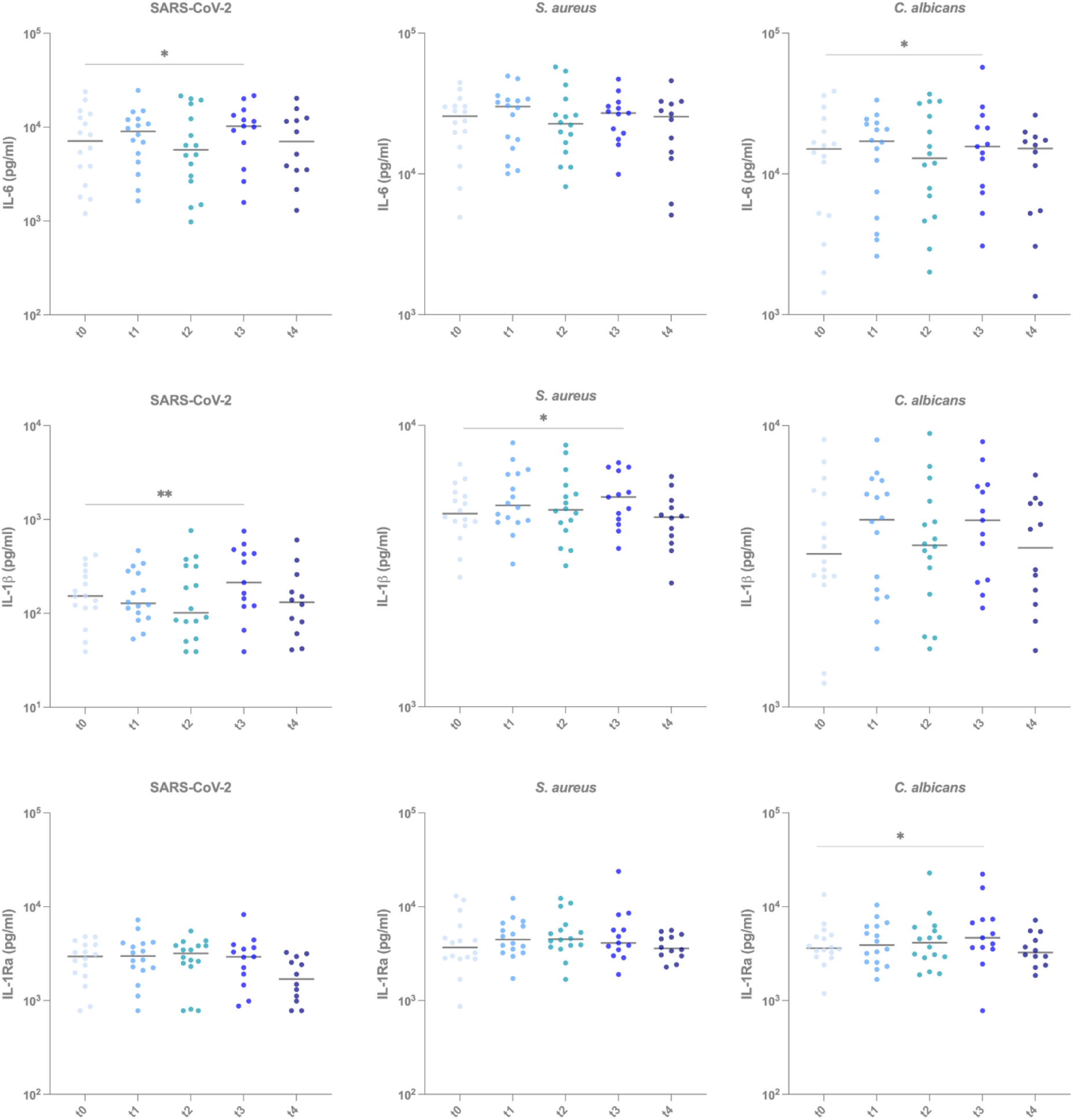
Inflammation-related cytokine production against viral, bacterial and fungal stimuli fluctuates through different time points. IL-6, IL-1β, and IL-1Ra production by human PBMCs measured by ELISA following 24 hours *ex vivo* stimulation with heat-killed SARS-CoV-2, heat-killed *S. aureus* and heat-killed *C. albicans at* t0 (before vaccination), t1 (three weeks after the first dose), t2 (two weeks after the second dose), t3 (six months after the first dose) and t4 (four weeks after the booster vaccination, which was approximately one year after the first dose). Wilcoxon paired test (not corrected for multiple comparisons) is used to compare the cytokine values measured at different timepoints to the those of t0 (before vaccination). * p<0.05, ** p<0.01. IL-6, interluekin 6; IL-1β, interleuking 1 beta; IL-1Ra, interleukin-1 receptor antagonist.

Interferons are essential for antiviral immunity [10]. Therefore, we measured IFN-α, a type I interferon, in the PBMCs in response to viral stimuli (SARS-CoV-2, influenza, poly I:C, and R848). IFN-α release in response to SARS-CoV-2 decreased over time, with the highest production before the vaccination and the lowest after the booster shot (Fig 6a). We observed a similar drop in the release of IFN-α compared to baseline in response to the TLR3 ligand poly I:C and TLR7/8 ligand R848, potent inducers of IFN-α [11].

**Figure 6.**
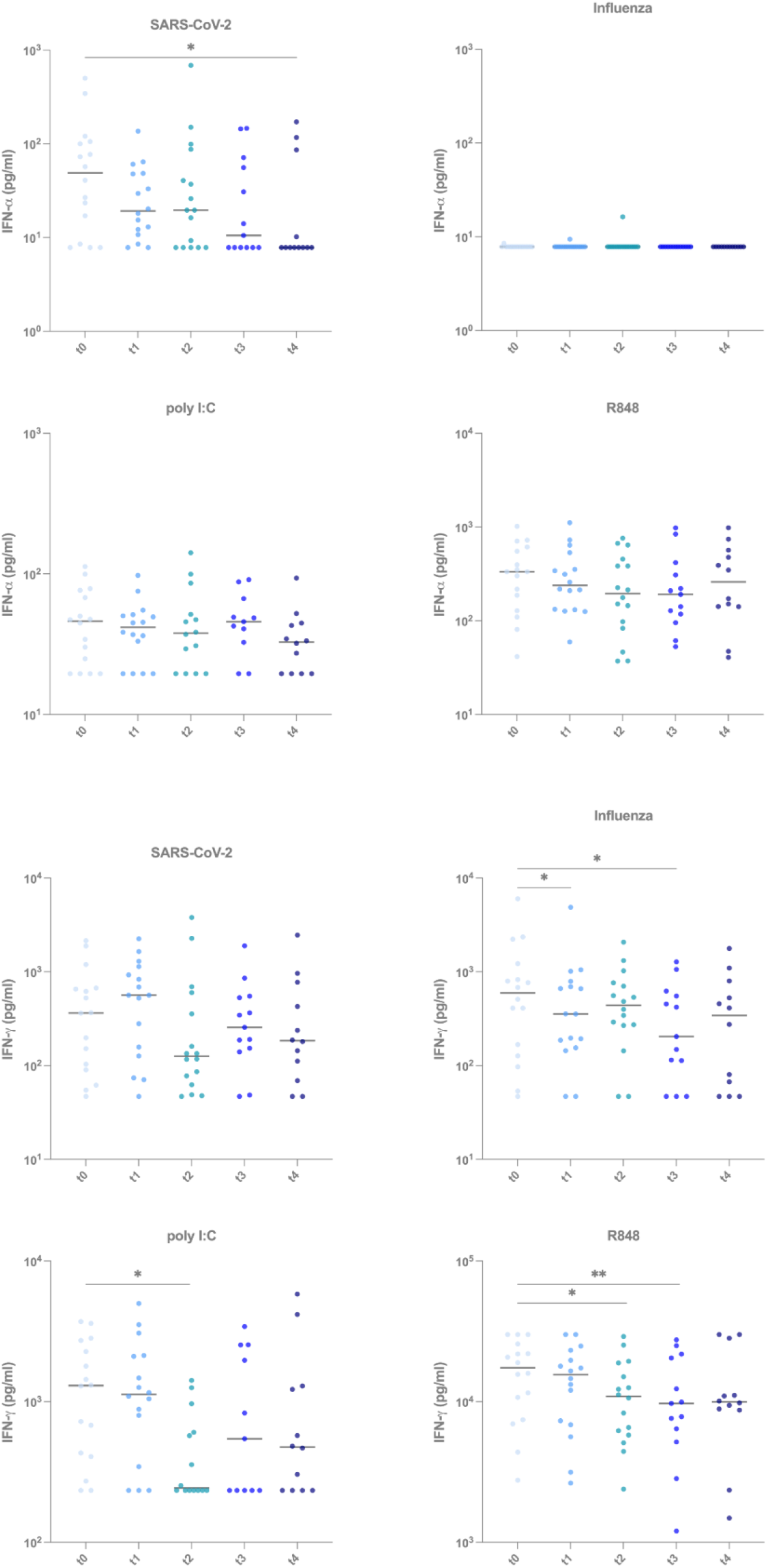
Type I and type II interferon production after *ex vivo* viral stimulation shows tendency to decrease after vaccination. **(a)** IFN-α measured by ELISA following 24 hours *ex vivo* stimulation in reponse to heat-killed SARS-CoV-2, heat-killed *S. aureus* and heat-killed *C. albicans at* t0 (before vaccination), t1 (three weeks after the first dose), t2 (two weeks after the second dose), t3 (six months after the first dose) and t4 (four weeks after the booster vaccination, which was approximately one year after the first dose). **(b)** IFN-γ measured by ELISA following 7 days of *ex vivo* stimulation with heat-killed SARS-CoV-2, heat-killed *S. aureus* and heat-killed *C. albicans at* t0, t1, t2, t3 and t4. Wilcoxon paired test (not corrected for multiple comparisons) is used to compare the cytokine values measured at different timepoints to the those of t0 (before vaccination). * p<0.05, ** p<0.01. IFN-α, interferon alpha; IFN-γ, interferon gamma.

Subsequently, we measured IFN-γ, a type II interferon produced mainly by NK and T cells, which showed a varying pattern after viral stimulation. There was a decrease in IFN-γ production after the second dose for all stimuli, reaching statistical significance for poly I:C and R848 (Fig 6b). Additionally, the cytokine production continuously dropped with each time point in the case of R848 stimulation. Especially for influenza, we observed a lower production of IFN-γ at t1 and t3 than at baseline.

## Discussion

Novel vaccines based on mRNA technology have been recently developed against COVID-19, but much remains to be learned about their wide immunological effects. In this study, we investigated the specific humoral effects of the BNT162b2 vaccine developed by BioNTech/Pfizer, as well as its effects on the innate immune responses to various viral, bacterial, and fungal pathogens. We show that the BNT162b2 vaccine induces long-term effects on both adaptive and innate immune responses, including transcriptional changes and effects on cytokine production capacity.

Our findings on the induction of specific anti-SARS-CoV-2 antibodies align with several recent studies. In this respect, we observed a decline in antibody concentrations and neutralizing capacity six months after vaccination [12,13]. It has been hypothesized that this decline is likely caused by plasmablasts that do not differentiate into long-lived memory plasma cells [14]. Booster vaccination restored the antibody concentrations, even higher than the concentrations after the first vaccination cycle. However, this increase was not significant, which differs from a few recent reports [15,16]. This discrepancy may be attributed to our reduced sample size, as reports from Salvagno and Teresa Vietri *et al*. have a 3-to 4-fold higher number of individuals included in their studies. In contrast, the neutralizing antibody titers did reach higher levels after the third than after the second vaccination.

The lack of neutralizing capacity against the omicron variant after six months underlines the urgent need for booster vaccinations that can target new variants, a finding also reported by others [17,18]. Omicron strains are, at the moment of writing, the most prevalent circulating variants, characterized by large numbers of mutations in the spike protein [19]. Those mutations, together with a recently detected higher affinity for the receptor angiotensin-converting enzyme 2 (ACE2), are suspected to be the reason why omicron variants so effectively escape antibody recognition [20].

In the last decade, an increasing number of studies have reported the long-term effects of vaccines not only on adaptive immune responses but also on innate immunity [21]. This results in a *de facto* innate immune memory termed *trained immunity*, which can result in protective effects against heterologous infections [22]. The newly developed COVID-19 vaccines, including those based on mRNA, were shown to have strong inflammatory effects due to their LNP delivery system [4], and we, therefore, set out to assess their potential long-term effect on the induction of trained immunity. First, we assessed the changes in the transcriptional program induced by viral stimulation with inactivated SARS-CoV-2, H1N1 influenza virus, or the TLR7/8 agonist R848 in immune cells after BNT162b2 vaccination. We observed that, after BNT162b2 vaccination, even unstimulated cells (RPMI) showed changes in the number of DEGs. A possible explanation for this might be the inflammatory feature of the LNP-delivered and modified-mRNA components of BNT162b2, as has been pointed out in a mouse model [4]. The multiple downregulated pathways observed following vaccination in unstimulated state may be caused by the exhaustion of the cells under the effects of such an inflammatory stimulus for a long time which mimics chronic infection. It is known that chronic inflammation can result in, for instance, T-cell exhaustion [23].

Recently, Arunachalam *et al*. showed that the second BNT162b2 vaccination generated a more pronounced transcriptional response as there was a general increase in the immune responsiveness [5]. However, the authors did not investigate the transcriptional response of immune cells post-vaccination upon viral restimulation. Our data underline the importance of investigating transcriptional responses after a perturbation rather than only in steady-state conditions. On the other hand, we observed increased numbers of DEGs at each consecutive time point in line with the work from Arunachalam *et al*., except those in SARS-CoV-2 stimulated cells, which decreased after secondary vaccination. Additionally, Yamaguchi *et al*. performed ATAC-sequencing of monocytes and observed an initial enhancement of type I IFN-related gene accessibility after the second vaccination, which disappeared after only four weeks [24]. In contrast, our study showed either no change or downregulation in the type I IFN-related pathways, which might suggest that BNT162b2 vaccination induces innate immune memory that also consists of tolerance characteristics.

Similar changes in the production of cytokines and interferons accompany the transcriptional responses to viral stimulation after vaccination. In this respect, the production of the cytokines from the IL-1/IL-6 pathway, including the anti-inflammatory IL-1Ra, tended to increase six months after the first vaccination. More remarkable is, however, the tendency of lower interferon responses after BNT162b2 vaccination. BNT162b2 vaccination has been previously reported to activate virus-specific CD4+ and CD8+ T cells and upregulate the production of immune-modulatory cytokines such as IFN-γ shortly after primary and secondary vaccination [5,25–27]. In contrast to these studies, we observed a downregulation of the type I interferon pathway in response to influenza at the transcriptional level, and lower IFN-α production by

PBMCs after stimulation with SARS-CoV-2. A similar pattern can be observed for IFN-γ, which was produced less by PBMCs after immunization when exposed to various viral stimuli. The cause of these different findings is unclear, although using different methodologies may partially explain them. Samanovic et. al., for instance, assessed the percentage of IFN-γ+ CD4+, and CD8+ T cells in response to the spike-peptide mix after a relatively short stimulation [25], whereas we measured the secreted cytokine in response to the heat-inactivated virus after a 7-days stimulation period.

To understand the heterologous effects of the BNT162b2 vaccine, one should also consider its composition. BNT162b2 comprises N1-methyl-pseudouridine (m1Ψ) nucleoside-modified mRNA encapsulated in a lipid nanoparticle (LNP) [28]. The pseudouridine modification increases mRNA stability and decreases an anti-RNA immune response [29]. LNPs are chosen as a delivery system for mRNA, which have been first conceptualized decades ago [30–32], and more recently, they have been used to deliver an RNA-based drug (Patisiran®) successfully [33]. Later on, Kariko *et al*. reported that infection of the cells with m1Ψ nucleoside-modified mRNA could dampen the response through TLR3 and TLR7 [29], which is in line with the results of our study. It could be hypothesized that this is due to the decrease in sensitivity of endosomal TLRs that interact with this transfected modified mRNA, subsequently ablating the activity of TLR3, TLR7, and TLR8 and decreasing cytokine production [34].

Another contributing factor in the success of mRNA vaccines was suggested to be the presence of an immunoadjuvant [29,35]. It is known that BNT162b2 does not contain conventional adjuvants; however, LNPs have been shown to act as immunostimulatory adjuvants besides their role in the delivery of the mRNA [35–37]. These findings support the more recent report from Ndeupen et al., showing highly inflammatory characteristics of LNPs in a mouse model [4], which can contribute to the effects of BNT162b2 and other mRNA vaccines.

The results of the present study support the hypothesis that BNT162b2 has long-term heterologous effects on immune cells, reminiscent of the induction of trained immunity [22]. In a mouse model, Qin et al. showed that mRNA-LNP pre-exposure can alter immune responses to influenza and *C. albicans* infection [6]. These results collectively demonstrate that the effects of the BNT162b2 vaccine go beyond the adaptive immune system and can also modulate innate immune responses. An important question, however, relates to the biological consequences of these effects. Recently, some debate has been ongoing on whether BNT162b2 immunization is associated with the reactivation of varicella-zoster virus (VZV) [38]. Possible explanations include the suppression of VZV-specific CD8+ cells by the immense shift of naïve CD8+ cells after immunization and the downregulation of TLR pathways through immunization and thereby inhibiting of interferon production. The hypothesis of downregulated interferon production agrees with our data, however, the causes of these effect remain to be proven. Our results may explain the data from a recent study of over 50,000 healthcare workers that found that the more doses of the mRNA vaccine received by the individuals, the higher their risk of contracting COVID-19 [39]. It may be thus hypothesized that vaccination with mRNA-based vaccines causes dysregulation of innate immune responses, and that the consequences of this effect for protection against SARS-CoV-2 cannot be fully compensated by the induction of adaptive immune responses.

On the other hand, the more dampened transcriptional reactivity of the immune cells to secondary viral stimulation (immune tolerance) may provide an explanation for the protective effects of BNT162b2 against severe COVID-19. Overwhelming inflammation is one of the important pathological features in patients with COVID-19. Thus, a more regulated inflammatory response may explain why vaccination had especially effects on the reduction of disease severity in case of the delta and omicron variants, rather than a full protection against infection [40]. Indeed, the complete absence of neutralization capacity against omicron in this and other studies argues that cellular mechanisms, such as other T-cell-mediated or innate immune cell-mediated pathways, are responsible for these effects. Furthermore, downregulation of type I IFN signaling has been suggested to be one of the mechanisms affecting immune memory to SARS-CoV-2 as the generation of long-lived memory cells is dependent on it [41,42].

The generalizability of these results is subject to certain limitations. First, the number of volunteers in this study was relatively small, although in line with earlier immunological studies on the effects of COVID-19 vaccines. Second, our cohort consisted of healthcare workers, who are middle-aged and healthy, and future studies on elderly individuals and people with comorbidities and other underlying risk factors for severe COVID-19 infections need to be performed. Third, our study is performed only with individuals with Western European ancestry. Therefore, the conclusions of our study should be tested in populations with different ancestry and alternative lifestyles since the induction of innate and adaptive immune responses is mainly dependent on factors such as genetic background, diet, and exposure to environmental stimuli, which differ between communities around the globe.

In conclusion, our data show that the BNT162b2 vaccine induces effects on both the adaptive and innate branches of the immune system. Intriguingly, the BNT162b2 vaccine induces significant changes in interferon production, and this needs to be studied in more detail: in combination with strong adaptive immune responses, this could contribute to a more balanced inflammatory reaction during infection with SARS-CoV-2 or other pathogens. Our findings need to be confirmed by conducting larger cohort studies with populations with diverse backgrounds, while further studies should investigate the incidence of heterologous infections after BNT162b2.

## Materials and Methods

### Cohort

Healthcare workers from the Radboud University Medical Center, Nijmegen were enrolled who received the BNT162b2 mRNA COVID-19 vaccine as per national vaccination campaign and provided informed consent. Key exclusion criteria included a medical history of COVID-19. Participants were asked at each study visit whether they had COVID-19 since the previous study visit. One individual was removed from the dataset after detecting high concentrations of antibodies against SARS-CoV-2 N-antigen at baseline and two individuals were removed at later time points because they had COVID-19 in the course of the study.

### Virus isolation and sequencing

Viruses were isolated from diagnostic specimen at the department of Viroscience, Erasmus MC, and subsequently sequenced to rule out additional mutations in the S protein. SARS-CoV-2 isolate BetaCoV/Munich/BavPat1/2020 (European Virus Archive 026V-03883), was kindly provided by Prof. C. Drosten. At 72 h post-infection, the culture supernatant was centrifuged for 5 min at 1500 x g and filtered through an 0.45 μM low protein binding filter (Sigma-Aldrich, Germany, cat #SLHPR33RS). To further purify the viral stocks, the medium was transferred over an Amicon Ultra-15 column with 100 kDa cutoff (Sigma-Aldrich, Germany, cat #UFC910008), which was washed 3 times using Opti-MEM supplemented with GlutaMAX (Thermo Fisher Scientific, USA, cat #51985034). Afterwards, the concentrated virus on the filter was diluted back to the original volume using Opti-MEM, and the purified viral aliquots were stored at -80 °C. The infectious viral titers were measured using plaque assays as described [43] and stocks were heat inactivated for 60 min at 56 °C for use in stimulation experiments.

### Measurement of antibody concentrations against RBD and Spike protein

For antibody analysis, a fluorescent-bead-based multiplex immunoassay (MIA) was developed, as previously described by Fröberg et al., 2021, with some slight modifications [44]. The first international standard for anti-SARS-CoV-2 immunoglobulin, (20/136, NIBSC), was used to create standard curves. Next to this, four different samples from PCR-confirmed COVID-19 patients were used as quality control samples. Serum samples were diluted 1:500 and 1:8000 in assay buffer (SM01/1%BSA), and incubated with antigen-coated microspheres for 45 min at room temperature while shaking at 450 rpm. Purified S (Stabilized Trimeric Spike Protein from the Wuhan variant) and receptor binding domain (RBD from the Wuhan variant) proteins purchased from ExcellGene were coupled to microspheres. Following incubation with sera, the microspheres were washed three times with PBS/0,05% Tween-20, incubated with phycoerythrin-conjugated goat anti-human, IgG (Jackson Immunoresearch, 109-116-170) for 20 min and washed three times. Data were acquired on the Luminex FlexMap3D System. Validation of the detection antibodies was obtained from a recent publication using the same antibodies and the same assay [45], and specificity was checked using rabbit anti-SARS SIA-ST serum. MFI was converted to International Units (IU/ml) by interpolation from a log-5PL-parameter logistic standard curve and log–log axis transformation using Bioplex Manager 6.2 (Bio-Rad Laboratories) software and exported to R-studio.

### Plaque reduction neutralization assay

A plaque reduction neutralization test (PRNT) was performed. Viruses used in the assay were isolated from diagnostic specimen at the department of Viroscience, Erasmus MC, cultured and subsequently sequenced to rule out additional mutations in the S protein. Heat-inactivated sera were 2-fold diluted in Dulbecco modified Eagle medium supplemented with NaHCO3, HEPES buffer, penicillin, streptomycin, and 1% fetal bovine serum, starting at a dilution of 1:10 in 60 μL. We then added 60 μL of virus suspension (400 plaque-forming units) to each well and incubated at 37 °C for 1h. After 1 hour incubation, we transferred the mixtures on to Vero-E6 cells and incubated for 8 hours. After incubation, we fixed the cells with 10% formaldehyde and stained the cells with polyclonal rabbit anti-SARS-CoV antibody (Sino Biological) and a secondary peroxidase-labeled goat anti-rabbit IgG (Dako). We developed signal by using a precipitate forming 3,3′,5,5′-tetramethylbenzidine substrate (True Blue; Kirkegaard and Perry Laboratories) and counted the number of infected cells per well by using an ImmunoSpot Image Analyzer (CTL Europe GmbH). The serum neutralization titer is the reciprocal of the highest dilution resulting in an infection reduction of >50% (PRNT50). We considered a titer >20 to be positive based on assay validation.

### Isolation of peripheral blood mononuclear cells

Blood samples from participants were collected into EDTA-coated tubes (BD Bioscience, USA) and used as the source of peripheral blood mononuclear cells (PBMCs) after sampling sera from each individual. Blood was diluted 1:1 with PBS (1X) without Ca++, Mg++ (Westburg, The Netherlands, cat #LO BE17-516F) and PBMCs were isolated via density gradient centrifuge using Ficoll-PaqueTM-plus (VWR, The Netherlands, cat #17-1440-03P).

Specialized SepMate-50 tubes were used for the isolation (Stem Cell Technologies, cat #85450). Cells counts were determined via Sysmex XN-450 (Japan) hematology analyzer. Afterwards, PBMCs were frozen using Recovery Cell Culture Freezing Medium (Thermo Fisher Scientific, USA, cat #12648010) in the concentration of 15×10^6^/mL.

### Stimulation experiments

The PBMCs were thawed and washed with 10mL Dutch modified RPMI 1640 medium (Roswell Park Memorial Institute; Invitrogen, USA, cat # 22409031) containing 50 μg/mL Gentamicine (Centrafarm, The Netherlands), 1 mM Sodium-Pyruvate (Thermo Fisher Scientific, USA, cat #11360088), 2 mM Glutamax (Thermo Fisher Scientific, USA, cat #35050087) supplemented with 10% Bovine Calf Serum (Fisher Scientific, USA, cat #11551831) twice. DNase (Roche, Switzerland, cat #1128493200) was added to the wash medium to digest extracellular DNA released from dying cells. Afterwards, the cells were counted via Sysmex XN-450. PBMCs (4×105 cells/well) stimulated in sterile round bottom 96-well tissue culture treated plates (VWR, The Netherlands, cat #734-2184) in Dutch modified RPMI 1640 medium containing 50 μg/mL Gentamicine, 1 mM Sodium-Pyruvate, 2 mM Glutamax supplemented with 10% human pooled serum. Stimulations were done with heat-inactivated SARS-CoV-2, Wuhan Hu-1 (GISAID accession number is EPI_ISL_425126, Wuhan-Hu-1 WT [46], (2.8×10^3^ TCID50/mL), influenza virus reference strain A/California/7/2009 H1N1 was used (described in [47]) (3.6×10^3^ TCID50/mL), 10 μg/mL Poly I:C (Invivogen, USA, cat #tlrl-pic), 3 μg/mL R848 (Invivogen, USA, cat #tlrl-r848), 1 × 10^6^/mL S. aureus and 1 × 10^6^ /mL C. albicans. The PBMCs were incubated with the stimulants for 24 hours to detect IL-1β, TNF-α, IL-6, IL-1Ra and IFN-α, and 7 days to detect IFN-γ as well as bulk RNA isolation. Supernatants were collected and stored in -20 °C. Secreted cytokine levels from supernatants were quantified by ELISA (IL-1β cat # DLB50, TNF-α cat # STA00D, IL-6 cat # D6050, IL-1Ra cat # DRA00B, IFN-γ cat #DY285B, R&D Systems, USA and IFN-α cat #3425-1H-20, Mabtech, Sweden) following manufacturers’ instructions.

### Bulk RNAseq analysis

Bulk RNA sequencing data were processed using the publicly available nfcore/rnaseq pipeline (v2.0, [48]), implemented in Nextflow (v21.04.3, [49]) using default settings. Reads were aligned to the human GRCh38 genome.

Further downstream analyses were performed in R (v4.2.0, [50] using DESeq2 (v1.36, [51]). Manual curation of quality control metrics and principal component analysis (PCA) was performed per timepoint and stimulation separately to identify potential outliers. 13 out of 242 samples were removed from further analysis.

### Statistical analysis

Graphpad Prism 8 was used for all statistical analyses. Outcomes between paired groups were analyzed by Wilcoxon’s matched-pairs signed-rank test (not corrected for multiple comparisons). Three or more groups were compared using Kruskal-Wallis Test -Dunnet’s multiple comparison. A p-value of less than 0.05 was considered statistically significant (* p<0.05, ** p<0.01, *** p<0.001, **** p<0.0001). Spearman correlation was used to determine correlation between groups.

For bulk RNA sequencing, differential gene expression was estimated using linear models that included individual’s sex and their age class (<40 or >40 years of age). Genes with total read counts < 20 were removed prior to this analysis. Resulting p-values were corrected across all genes using Benjamini-Hochberg. Genes with adjusted p-values < 0.05 and logFC > 0.5 were considered significantly differentially expressed. For gene set enrichment analysis, we used blood transcription modules [52]. Genes were ranked by the Wald statistic and p-values were adjusted using Benjamini-Hochberg. Adjusted P-values < 0.05 were considered significant.

### Study approval

The study was approved by the Arnhem-Nijmegen Institutional Review Board (protocol NL76421.091.21) and registered in the EU clinical trials register (EudraCT: 2021-000182-33).

## Data Availability

The data is available upon request.

## Data and code availability

The RNAseq datasets generated during and/or analyzed during the current study are included as electronic supplementary material. The other data is available upon request.

## Acknowledgements

We thank all the volunteers to the study for their willingness to participate and the research technicians (Helga Dijkstra, Heidi Lemmers, S. Andrei Sarlea and Maartje Reijnders) for their help in collecting samples.

## Declaration of Interests

M.G.N and L.A.B.J are scientific founders of TTxD and Lemba.

## Funding

L.A.B.J. is supported by a Competitiveness Operational Program Grant of the Romanian Ministry of European Funds (HINT, ID P_37_762; MySMIS 103587). J.D-A. is supported by the Netherlands Organization for Scientific Research (VENI grant 09150161910024 and Off-Road grant 04510012010022). M.G.N. is supported by an ERC Advanced Grant (#833247) and a Spinoza Grant of the Netherlands Organization for Scientific Research.

## Notes

### Clinical Trial

EudraCT 2021-000182-33

### Author Declarations

Arnhem-Nijmegen Institutional Review Board

### Summary of Updates

We have added follow-up data up to one year after the first administration of BNT162b2 and we also added to the manuscript data on transcriptional activity in the immune cells.

