## Supplementary figures and images for "The impact of BNT162b2 mRNA vaccine on adaptive and innate immune responses"

### Supplemental Figure 1

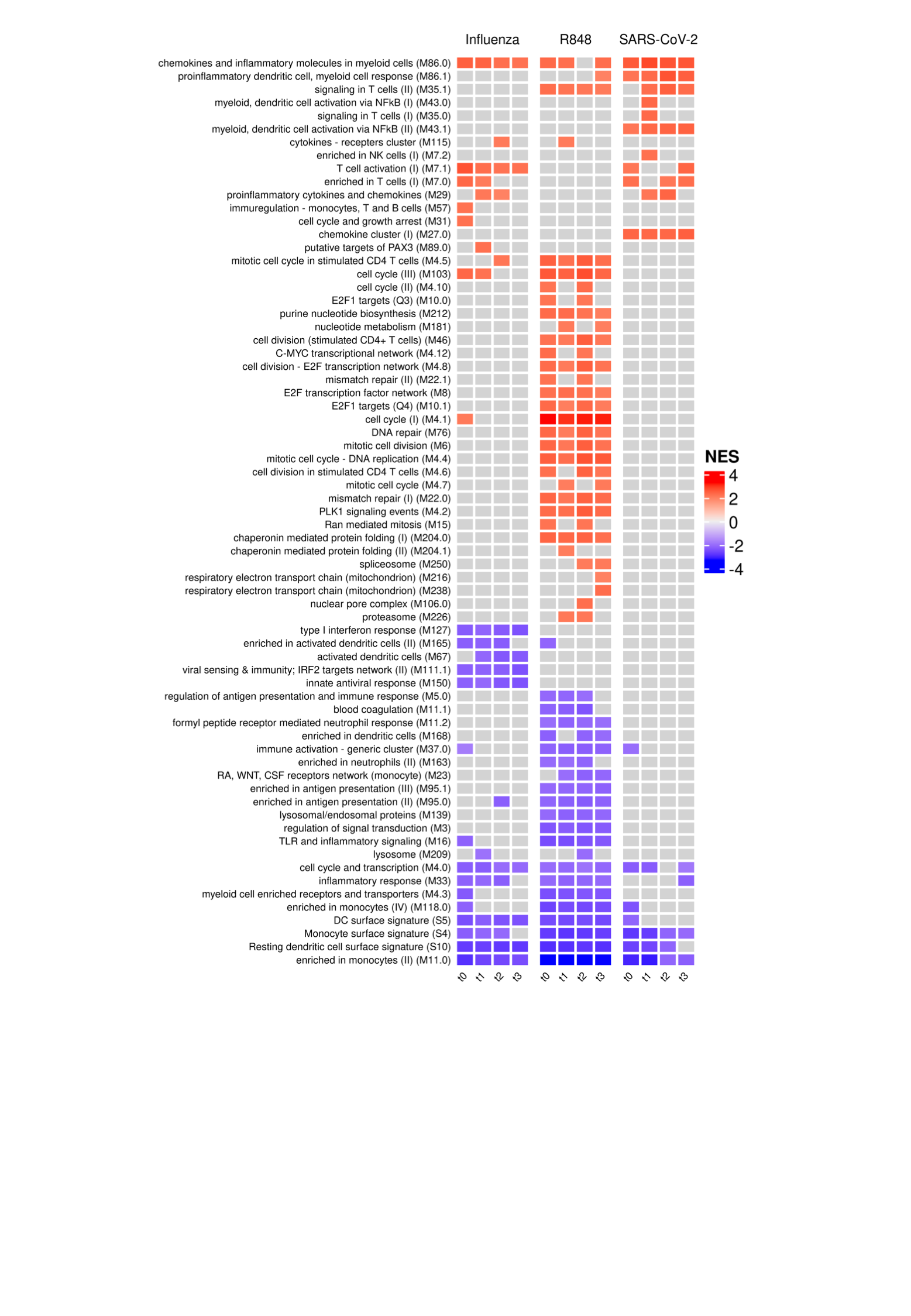

### Supplemental Figure 2

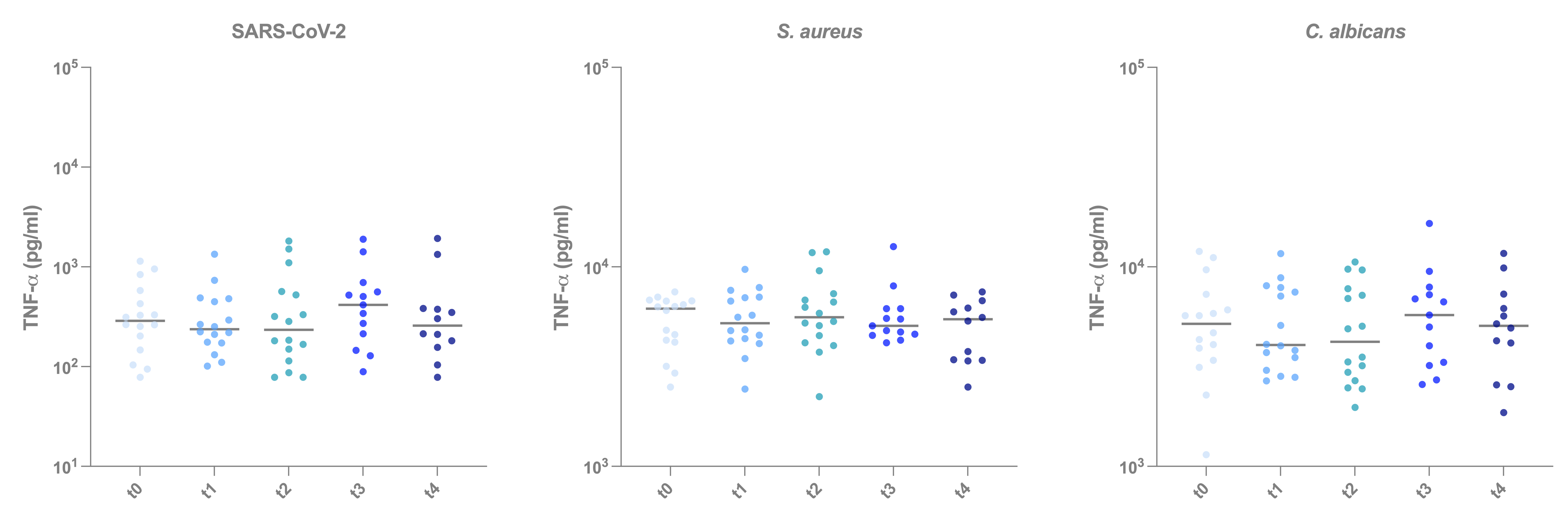
